# Proteomics Uncovers Cryptic *JPH2* Loss in Paediatric Dilated Cardiomyopathy

**DOI:** 10.64898/2026.06.16.26355718

**Authors:** Carlos C. Smith-Diaz, Valerii Iaprintsev, Hannah Huckstep, Ivan Macciocca, Adam T. Piers, Natasha Henden, Samantha Bryen, Natalie Stewart, Alexandra Butters, Amy M. Baker, Laura Catto, Leah Kemp, Ingrid King, Lina H. H. Le, David A. Elliott, Kevin I. Watt, Jacob Mathew, Robert Justo, Ebony Richardson, Cas Simons, Andrew P. Landstrom, Christina V. Theodoris, Ira W. Deveson, Daniel G. MacArthur, Igor E. Konstantinov, Robert G. Weintraub, Enzo R. Porrello, Sean J. Humphrey, Jodie Ingles

**Affiliations:** Garvan Institute of Medical Research, and the University of New South Wales, Sydney, NSW, Australia; Department of Cardiac Surgery, The Royal Children’s Hospital, Melbourne, VIC, Australia; Department of Paediatrics, University of Melbourne, Melbourne, VIC, Australia; Murdoch Children’s Research Institute, Melbourne, VIC, Australia; Novo Nordisk Foundation Center for Stem Cell Medicine, Murdoch Children’s Research Institute, Melbourne, VIC, Australia; Victorian Clinical Genetics Services, Royal Children’s Hospital, Melbourne, VIC, Australia; Melbourne Centre for Cardiovascular Genomics and Regenerative Medicine, Melbourne, VIC, Australia; Centre for Population Genomics, Murdoch Children’s Research Institute, Melbourne, VIC, Australia; Centre for Population Genomics, Garvan Institute of Medical Research and the University of New South Wales, Sydney, NSW, Australia; Department of Anatomy & Physiology, School of Biomedical Sciences, The University of Melbourne, Melbourne, VIC, Australia; Department of Cardiology, The Royal Children’s Hospital, Melbourne, VIC, Australia; Queensland Children’s Hospital, Brisbane, QLD, Australia; Department of Pediatrics, The Children’s Hospital of Philadelphia, Philadelphia, PA, USA; Department of Pediatrics, Perelman School of Medicine, University of Pennsylvania, Philadelphia, PA, USA; Gladstone Institute of Cardiovascular Disease; Gladstone Institute of Data Science and Biotechnology, San Francisco, CA, USA; Department of Pediatrics, Institute for Human Genetics, Cardiovascular Research Institute, University of California, San Francisco, San Francisco, CA, USA; Faculty of Medicine and Health, University of New South Wales, Sydney, NSW, Australia

**Keywords:** Proteomics, paediatric cardiomyopathy, Junctophilin-2

## Abstract

Despite recent advances in next-generation sequencing, genetic diagnostic rates for dilated cardiomyopathy (DCM) remain low. Among paediatric DCM, causes are often heritable, with a greater frequency of *de novo*, recessive and syndromic causes of disease. Novel diagnostic methods are therefore required to solve monogenic cases. To assess the value of proteomics as a diagnostic tool for paediatric DCM, we obtained left ventricle myocardial samples from paediatric patients undergoing heart transplantation at the Royal Children’s Hospital, Melbourne. We performed genome sequencing and proteomics and leveraged this multi-omics dataset to uncover the molecular cause of disease in a gene elusive proband. The proband carried a heterozygous *JPH2* frameshift variant identified on clinical exome sequencing. However, proteomic analysis showed a pronounced downregulation of JPH2, suggestive of biallelic loss-of-function. Closer inspection of the genomic data revealed a large inversion (∼8.34 Mb) with a breakpoint falling within intron 5 of *JPH2* that displaces the 3′UTR from the coding transcript. The two variants were confirmed to be *in trans* using long read DNA sequencing, consistent with a diagnosis of *JPH2* autosomal recessive DCM. Finally, we applied RNA sequencing with total RNA library preparation to show that transcripts containing a 3′UTR were reduced to ∼10% relative to controls. As a proof-of-principle, we present the first reported use of proteomics from explanted cardiac tissue to provide a genetic diagnosis. Our methodology has broad relevance to patients with genetically unsolved Mendelian diseases, who might undergo organ transplantation as part of clinical management.

## INTRODUCTION

Junctophilin-2 (*JPH2*, HGNC:14202) is a vital component of the junctional membrane complex involved in excitation-contraction coupling in cardiomyocytes. JPH2 physically tethers the transverse tubules of the plasma membrane to the sarcoplasmic reticulum, and is therefore critical for Ca2+-induced Ca2+ release following cardiomyocyte excitation^1,2^. Homozygous or compound heterozygous loss-of-function variants in *JPH2* cause a rare form of autosomal recessive dilated cardiomyopathy (DCM) characterised by a severe early-onset phenotype^3–6^. The ClinGen Gene Curation working group assesses the current evidence level for a gene-disease association, and defines a strong level of evidence for biallelic variants in *JPH2* and DCM^7^. In contrast, heterozygous *JPH2* missense variants cause hypertrophic cardiomyopathy (HCM) via a dominant-negative mechanism^6^, with a moderate gene-disease association^8^.

Here, we illustrate the value of biobanking explanted myocardium for diagnostic proteomics. We highlight the utility of our methodology by showing how this allowed us to provide a genetic diagnosis to a patient with paediatric DCM requiring heart transplantation. The proband was gene elusive following clinical exome sequencing. Using a multi-omics diagnostic approach, involving proteomics, RNA sequencing, short-read and long-read genome sequencing, we show that biallelic *JPH2* loss-of-function underpins the proband’s phenotype. Our work provides a proof-of-principle example of the value of proteomics using explanted cardiac tissue to provide a genetic diagnosis to a patient with DCM.

## METHODS

### Family Recruitment

The family provided written informed consent for the Melbourne Children’s Heart Tissue Bank (HREC 38192) which included storage of cardiac transplant tissue for proteomics analysis. Following inconclusive clinical exome sequencing, the proband and her parents were recruited to Elusive Hearts, an Australian study aiming to identify novel causes of monogenic cardiovascular disease. The Elusive Hearts study received ethics approval from the Royal Children’s Hospital Human Research Ethics Committee, #97521. Participants provided written informed consent. Medical records were obtained, and a detailed family history was collected.

### Genome Sequencing, Alignment and Variant Calling

Library preparation for genome sequencing was performed from granulocyte-extracted DNA from the proband using the Illumina DNA Prep PCR-free library preparation kit, and sequenced by the Australian Genome Research Facility (AGRF) on an Illumina NovaSeq 6000 platform to achieve a target yield of 100 Gbp raw data.

Genome sequencing data processing was performed using the Centre for Population Genomics (CPG) CaRDinal platform following the DRAGEN GATK (Genome Analysis Toolkit) best practices. Reads were aligned to the hg38 reference genome using Dragmap (v1.3.0). Cohort-wide joint calling of single nucleotide variants (SNVs) and small insertion/deletion (indel) variants was performed using GATK HaplotypeCaller (v4.2.6.1) with “--dragen-mode” enabled. Variants were annotated using VEP 110. Structural variant (SV) calling was performed using GATK-SV^9^. The resulting callset was loaded into CPG’s CaRDinal deployment of *seqr* for analysis^10^. Mitochondrial variants were called from genome sequencing data using the GATK mitochondrial variant calling workflow with MuTect2 in mitochondria mode^11^, and outputs were processed to generate MitoReports for downstream review. Sample sex and relatedness quality checks were performed using Somalier (v0.2.15)^12^.

### Variant Filtering and Analysis

We analysed a virtual panel of 656 cardiac genes, created by merging all green, amber and red genes listed in at least one of the following panels in the PanelApp Australia database on 19 January 2026: Adult Cardiac Super Panel v2.112, Arrhythmia Super Panel v3.72, Cardiomyopathy Adult Super Panel v2.72, Cardiomyopathy Paediatric Panel v0.217, Congenital Heart Defect Panel v0.521, Aortopathy Connective Tissues Disorders Panel v1.101, Familial Hypercholesterolaemia Panel v1.0, Spontaneous Coronary Artery Dissection Panel v0.56 and/or Pulmonary Arterial Hypertension Panel v1.50.

Candidate variant analysis was restricted to rare variants (<0.0001 mean allele frequency [MAF] in gnomAD v2.1.1 or v3.1.2 for SNVs). All rare SNVs present in at least one allele in the affected proband; and with an Ensembl Variant Effect Prediction (VEP) of high, moderate or low impact, or with a SpliceAI maximum raw delta score >0.10, were manually reviewed. All rare SVs were manually reviewed. Rare SVs were defined as those where SVs of the same type and with a sufficient reciprocal overlap (>10% for SVs <5kb, and >50% for SVs >5kb), or with a breakpoint within +/- 100bp for insertions, were present at <0.0001 MAF in gnomAD SVs v4.1.0; and at <0.001 MAF in the joint-called SV VCF for all genomes in the CPG-instance of *seqr* by 2 February 2026.

### Myocardial Tissue Acquisition

Myocardial tissues from children with DCM were collected from patients undergoing heart transplantation for end-stage heart failure at the Royal Children’s Hospital, Melbourne (RCH). All patients undergoing transplantation between 2017 and 2023 were included. At cardiac transplantation (n = 24), samples were obtained from the apical part of the left ventricle (LV). Healthy donor myocardial samples were obtained by endomyocardial excision from the same region in the LV (n = 14). After collection, samples were immediately washed in cold Plasma-Lyte solution, transported from the operating room on ice, and stored at −80°C within 60 minutes of acquisition.

### Proteomics Sample Preparation and Data Acquisition

Upon retrieval from the tissue bank, samples were maintained frozen on dry ice and pulverised into a fine powder under liquid nitrogen using a pre-chilled mortar and pestle. The powdered tissue was transferred into pre-chilled microcentrifuge tubes. Protein extraction was performed using a freshly prepared sodium deoxycholate (SDC)-based lysis buffer containing 4% SDC and 100 mM Tris-HCl (pH 8.5) in Milli-Q water. Protease and phosphatase inhibitors were not added, as samples were boiled immediately after lysis at 95°C for 5 minutes to ensure rapid inactivation of endogenous enzymes. For initial protein yield estimation, a representative aliquot of tissue powder was lysed in 100 µL of buffer, cooled on ice after heating, and sonicated using a tip-probe sonicator (50–70% output, 10-second pulses, 60 seconds total). Lysates were then vortexed thoroughly, and a 10 µL aliquot was diluted 1:5 in 8 M urea for protein quantification using the bicinchoninic acid assay. Based on the measured protein yield, larger tissue aliquots were lysed using adjusted buffer volumes to achieve a final protein concentration of 4 mg/mL. All samples were normalised to equal protein concentrations, with a minimum concentration of 4 mg/mL and a maximum working volume of 200 µL. Prepared lysates were stored at −80°C until mass spectrometry processing.

### Proteomic Bioinformatics Analysis

Proteomics data were processed and quantified using MaxQuant. The proteinGroups.txt output was exported into R for data cleaning and quality control using a custom script. Filtering was performed to remove potential contaminants. Protein intensities were normalised with a log2 transformation with median centering, and low abundance proteins were removed. Post-normalisation quality control was performed by generating PCA plots and density plots of log2 intensities post-normalisation.

We filtered the cleaned dataset for proteins where decreased protein abundance is a driver of cardiomyopathy, either in the context of autosomal dominant, recessive, semi-dominant or X-linked inheritance. To exclude low confidence hits, protein fragments with low abundance, a mean log2 normalised expression < 1 in the healthy donor LV proteome cohort, were excluded. Proteomics data was available for the following cardiac genes of interest: *ALPK3, BAG3, CSRP3, DSC2, DSG2, JPH2, LAMP2, LMNA, MYBPC3, MYLK3, NEXN, PKP2, TTN* and *VCL*. An outlier analysis was performed to detect protein outliers in the proband’s proteome relative to the DCM control cohort mean.

To assess changes in JPH2 peptide-level abundance, the peptides.txt output from MaxQuant was filtered for unique mapping peptides to JPH2 (Q9BR39;Q9BR39-2) with filtering to remove peptides not quantified in any of the DCM control samples. The difference in LFQ (label-free quantification) intensity was then compared for peptides in the proband vs the mean of the DCM control cohort (n=23).

### Oxford Nanopore Targeted Long-Read Sequencing

DNA was extracted from right atrial tissue from the proband’s explanted heart using the PacBio PanDNA kit (PacBio, Cat# 102-260-000). Genomic DNA was sheared to ∼20–30 kb fragment size using the Diagenode Megaruptor 3 DNA shearing system (speed 29), treated with the Short-read Eliminator kit (PacBio, Cat# 102-208-300) to deplete fragments <10 kb. An ONT (Oxford Nanopore Technologies) library was prepared using a ligation prep (SQK-NBD114.24). The library was loaded on an ONT PromethION R10.4.1 flow cell (FLO-PRO114M) and sequenced on a PromethION 48 instrument. Readfish^13^ (v0.0.10dev2) was used to execute live target selection, enriching a custom target region covering chr20q, which includes the *JPH2* locus and candidate inversion.

Raw ONT sequencing data were converted to BLOW5 format^14^ and base-called with Buttery-eel^15^ (a wrapper to enable BLOW5 input for ONT’s Dorado basecaller), using the ‘super-accuracy’ model with 5mC methylation calling enabled. Base-called data was then processed using pipeface (https://github.com/leahkemp/pipeface) (v0.10.2), a NextFlow pipeline for long read sequencing data analysis. As relevant here, Pipeface executed minimap2^16^ (v2.28-r1209) for alignment, Deepvariant^17^ (v1.8.0) for calling small variants, sniffles2^18^ (v2.6.0) and cuteSV^19^ (v2.1.1) for calling SVs and WhatsHap^20^ (v2.3) for phasing. Data visualisation was performed using IGV (Integrative Genomics Viewer) (v2.19.5). The IGV display window was exported as an SVG for formatting and annotation in Inkscape (v1.4.3).

### RNA Sequencing

Right atrial tissue was obtained from the proband’s explanted heart, three hearts from donors who died of non-cardiac causes and the explanted heart of a patient with paediatric-onset DCM who had also undergone transplantation at RCH. The paediatric DCM control carried a predicted NMD-incompetent frameshift variant in *NEXN*: c.1918_1922del, p.(Tyr640ThrfsTer14) with no identified pathogenic variants in *JPH2*. Total RNA was extracted using the EZ2 RNA/miRNA kit. RNA integrity was measured using an Agilent TapeStation system with sample RIN (RNA integrity number) scores between 6.4 - 9.4, including a RIN of 8 for the proband’s sample. Library preparation was performed using the Ilumina stranded total RNA prep library preparation kit. Libraries were sequenced on an Element Biosciences AVITI system using the AVITI 2×150 cloudbreak medium output kit, generating 150bp paired end reads (∼100 million reads per transcriptome).

FASTQ files were trimmed with Trim Galore using the default parameters (Phred score cutoff 20). Alignment against GRCh38 was performed on the Gadi HPC system (NCI, Australia) using STAR (v2.7.11b) with the following settings: --twopassMode Basic, --outSAMtype BAM SortedByCoordinate, - -outSAMstrandField intronMotif. Sashimi plots and quantification of split exon-exon junction reads was performed in IGV (v2.19.5). The IGV display window was exported as an SVG for formatting and annotation in Inkscape (v1.4.3). Log2 FPM (Fragments per million) normalised read counts were calculated in Seqmonk (v1.49.1). Junction reads spanning the *JPH2* Exon5-3′UTR junction were quantified by calculating junction spanning split reads and applying the following normalisation:

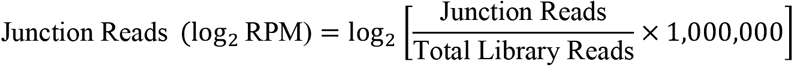

## RESULTS

A 10-15-year-old female was diagnosed with DCM after experiencing severe LV dysfunction and mild-moderate RV dysfunction. She developed cardiogenic shock with renal and liver dysfunction. She received a left ventricular assist device, with concurrent myocardial biopsy showing no signs of myocarditis, and subsequently underwent cardiac transplantation. She has remained in a stable condition several years post-transplant with good graft function and normal left ventricular function. There is no known family history of congenital or ischaemic heart disease. Both parents have had echocardiograms showing no evidence of DCM. The father has normal LV size and systolic function with a mildly dilated RV with normal systolic function. The mother has normal left and right ventricle size and systolic function.

The proband had clinical exome sequencing which revealed a likely pathogenic heterozygous frameshift variant in *JPH2:* NM_020433.5(*JPH2*): c.1359_1360insC, p.(Asp454ArgfsTer23). This variant induces a premature termination codon within exon 4 and is predicted to undergo nonsense mediated decay (NMD). The variant is rare, being present in three heterozygotes in gnomAD v4.1. No additional variants in *JPH2* were detected and no clinically significant abnormalities were observed on molecular karyotyping (including at the *JPH2* locus: 20q13.12). The frameshift variant was shown to have been inherited from the proband’s mother with Sanger sequencing. The case was ultimately considered genetically unsolved as truncating *JPH2* variants are not a recognised cause of autosomal dominant DCM^6^. *JPH2* is also relatively tolerant to loss-of-function variation (pLI=0, LOEUF=0.89, gnomAD v4.1.1) suggesting that heterozygous loss-of-function is unlikely to cause severe paediatric onset disease.

We performed a cardiac proteomics screen and observed a pronounced downregulation of JPH2 (UniProt ID: Q9BR39) in the proband’s left ventricle with a log2 fold change of -2.49 vs the transplant cohort mean and -2.60 vs the donor cohort mean (**Figure 1A**). The magnitude of change (∼5.6x decrease) was indicative of biallelic loss-of-function. We therefore performed a deep analysis of the genomic data for additional variants that might affect *JPH2* expression. We identified a ∼8.34 Mb inversion in chromosome 20 (located at chr 20:44114025-52454700), with the 5′ inversion breakpoint occurring within intron 5 of *JPH2*. Our manual review of aligned reads confirmed the location of these breakpoints (**Figure 1C**). While this inversion does not span the coding sequence of *JPH2*, it displaces the 3′UTR, which contains the poly-A signal sequence, from the coding sequence (**Figure 1B**). Detailed analysis of peptide-specific LFQ intensities showed that all peptides uniquely mapping to JPH2 were highly decreased in the proband relative to the DCM cohort mean, with relative peptide % abundances ranging from 0% to 28% and a mean peptide decrease in LFQ intensity ∼15.5% (**Figure 1D**).

**Figure 1:**
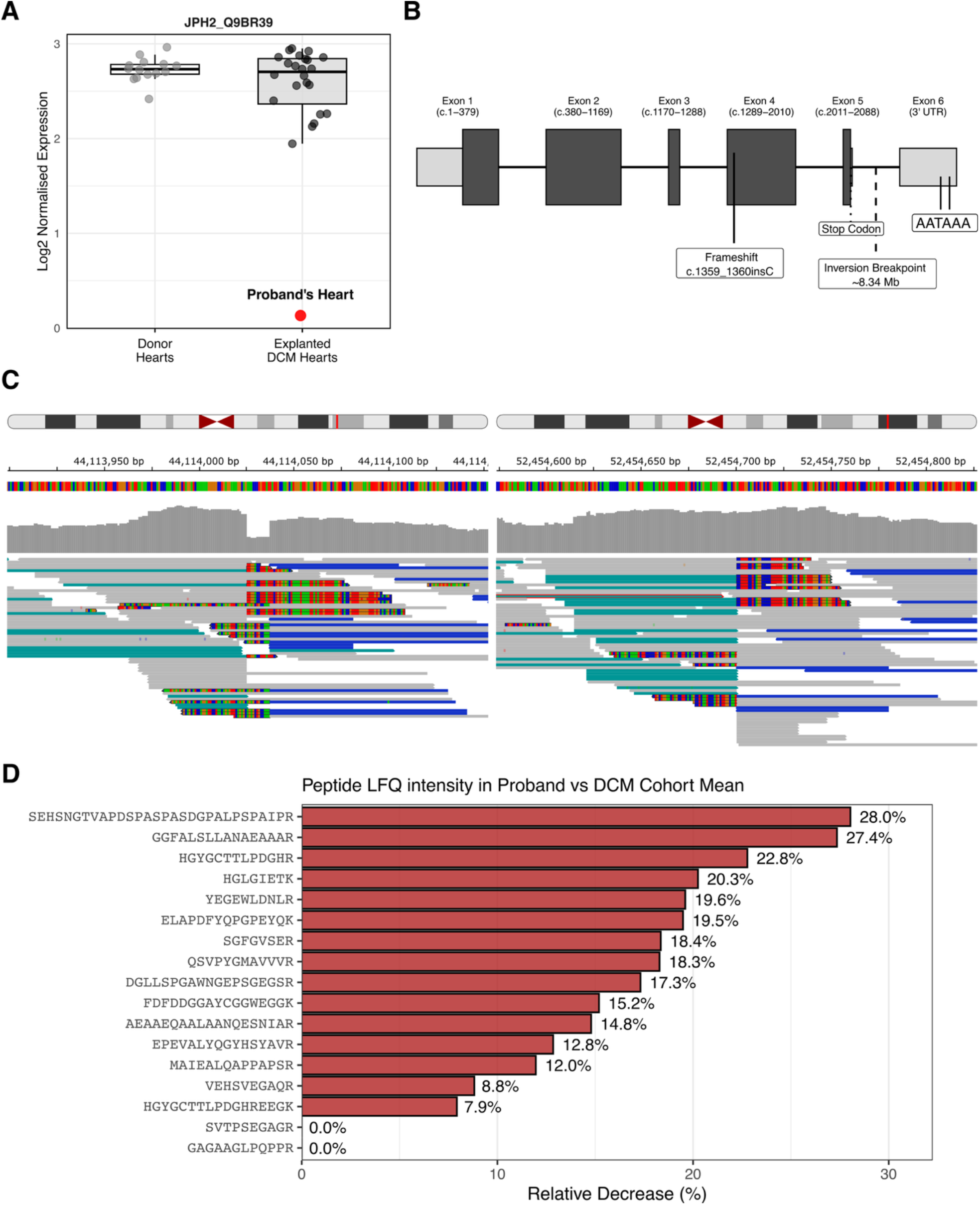
Proteomics reveals biallelic *JPH2* loss. **A:** JPH2 protein abundance in the proband’s LV relative to control LV tissue taken from healthy donor hearts (“Donor Hearts”) and explanted hearts from paediatric patients with end stage DCM (“Explanted DCM Hearts”). **B:** The positions of the frameshift allele (c.1359_1360insC, p.Asp454ArgfsTer23) and the inversion breakpoint depicted in the *JPH2* canonical transcript (ENST00000372980.4). Two poly-A signal motifs are located at the 3′ end of the 3′UTR. **C:** Standard short read genome sequencing data shown in the IGV window at positions chr20:44,113,899-44,114,152 and chr20:52,454,574-52,454,827 depicting the two inversion breakpoints at chr20:44,114,025 (*JPH2*) and chr20:52,454,700. Teal = read pairs mapping in the left-left orientation. Blue = read pairs mapping in the right-right orientation. **D:** Relative decrease in JPH2-peptide LFQ intensity in the proband’s heart relative to the mean JPH2-peptide LFQ intensity of the DCM cohort mean (n=23).

*JPH2* contains two poly-A signal sequence AATAAA motifs within its 3′UTR, one at chr20:44,108,127-44,108,132 and the other at chr20:44,107,065-44,107,070 (**Figure 1B**). The poly-A signal sequence is critical for mRNA 3′-end polyadenylation and variants affecting polyadenylation are known to cause Mendelian disorders^21,22^. Polyadenylation is required for mRNA export from the nucleus and, in general, transcripts lacking poly-A tails are very poorly translated^23^. The 3′UTR of *JPH2* may also have other important regulatory functions as it is a proposed binding site for the post-transcriptional regulator microRNA miR-34a^24^. Due to the displacement of the 3′UTR from the coding transcript, we predict that the resulting *JPH2* inversion transcript would lack a poly-A tail, and that transcribed mRNA would be subject to 3′-5′ exonucleolytic decay by the nuclear exosome complex^25^ or would otherwise be poorly translated due to export failure from the nucleus.

In addition to the ∼8.34 Mb inversion, we detected the heterozygous frameshift allele that had been reported with clinical exome sequencing. Variant phasing was performed with programmable targeted Oxford Nanopore long read sequencing, showing the inversion breakpoint and the frameshift allele on alternate haplotypes in the proband’s genome (**Figure 2**). The inversion was also detected using the sniffles2 structural variant caller from long read sequencing data. The proband’s phenotype of severe paediatric DCM was consistent with biallelic *JPH2* loss-of-function^3–5^, thus providing a molecular diagnosis.

**Figure 2:**
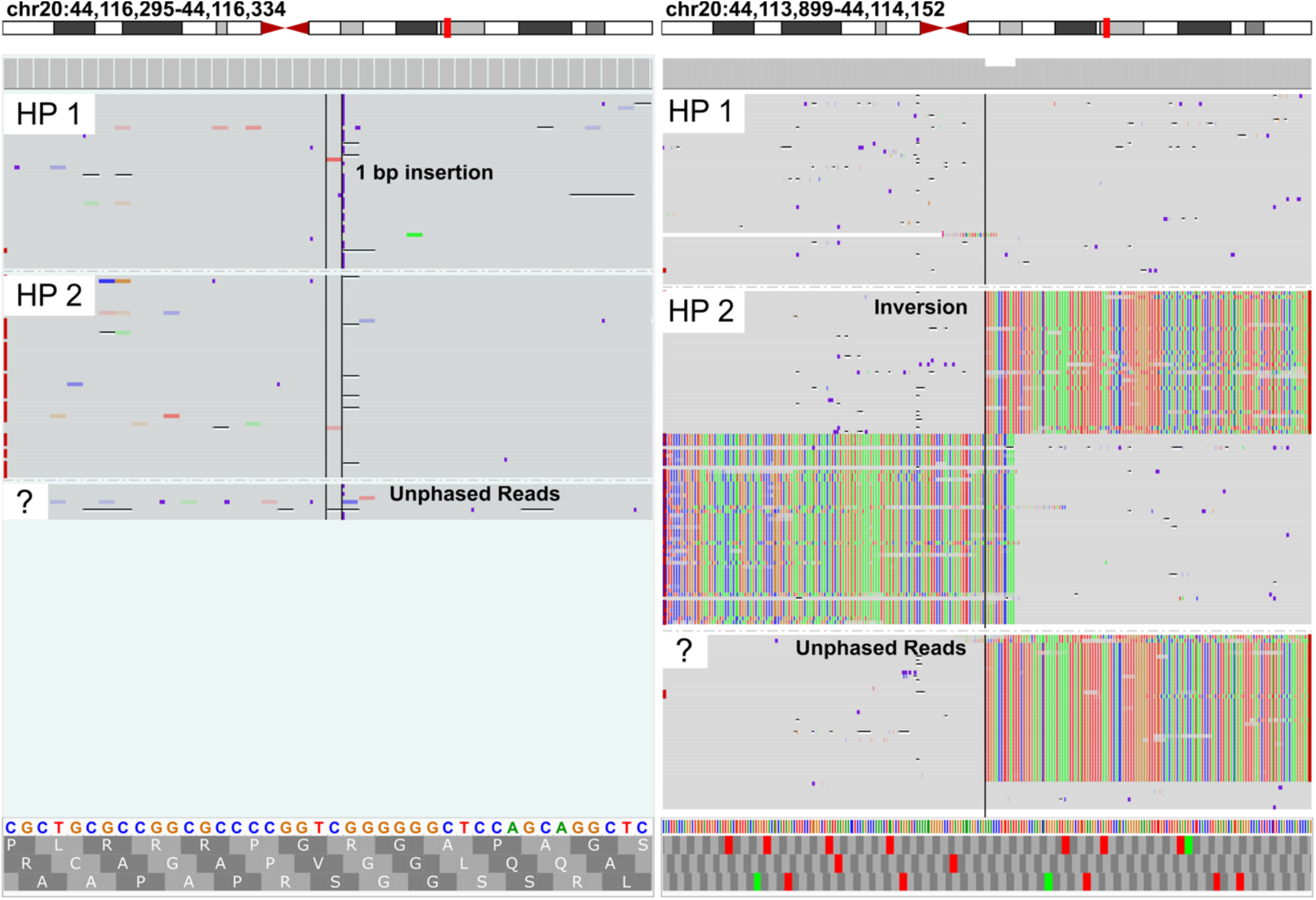
Variant phasing with long-read sequencing. Depiction of long-read genome sequencing data showing haplotypes 1 (HP 1) and 2 (HP 2) with the c.1359_1360insC variant observed on haplotype 1 at chr20:44,116,295-44,116,334 (*left*) and the inversion breakpoint observed on haplotype 2 within *JPH2* displayed at chr20:44,113,899-44,114,152 (*right*). Rainbow reads represent soft-clipped reads.

To exclude other potential genetic causes, we reviewed 656 cardiac disease-associated genes and did not identify any other variants that we considered causative of the proband’s phenotype. Further, we identified 48 genes present within the inverted region of chromosome 20 that were either dosage sensitive or had a definitive Mendelian disease association from ClinGen. 25 of these proteins were expressed within our cardiac proteomics dataset. For these 25 proteins, we observed no notable deviation in the proband’s proteome relative to the proteomes of other heart transplant patients.

Finally, we performed RNA sequencing using right atrial tissue from the proband’s heart. Total RNA library preparation with ribosomal depletion was performed based on the suspicion that poly-A enrichment would fail to capture non-polyadenylated transcripts from the inversion allele. Analysis of the RNA sequencing data showed a decrease in *JPH2* mRNA with a log2 fold change of -1.2 in the proband’s heart relative to controls (n=4) (**Figure 3A-B**). Moreover, when we considered the normalised abundance of junction reads spanning the 3′UTR-Exon 5 junction, quantified reads in the proband’s heart were more markedly reduced, down to ∼10% of the control mean. This suggests that very few *JPH2* transcripts would have a 3′UTR within the mature mRNA transcript in the proband’s heart (**Figure 3C**).

**Figure 3:**
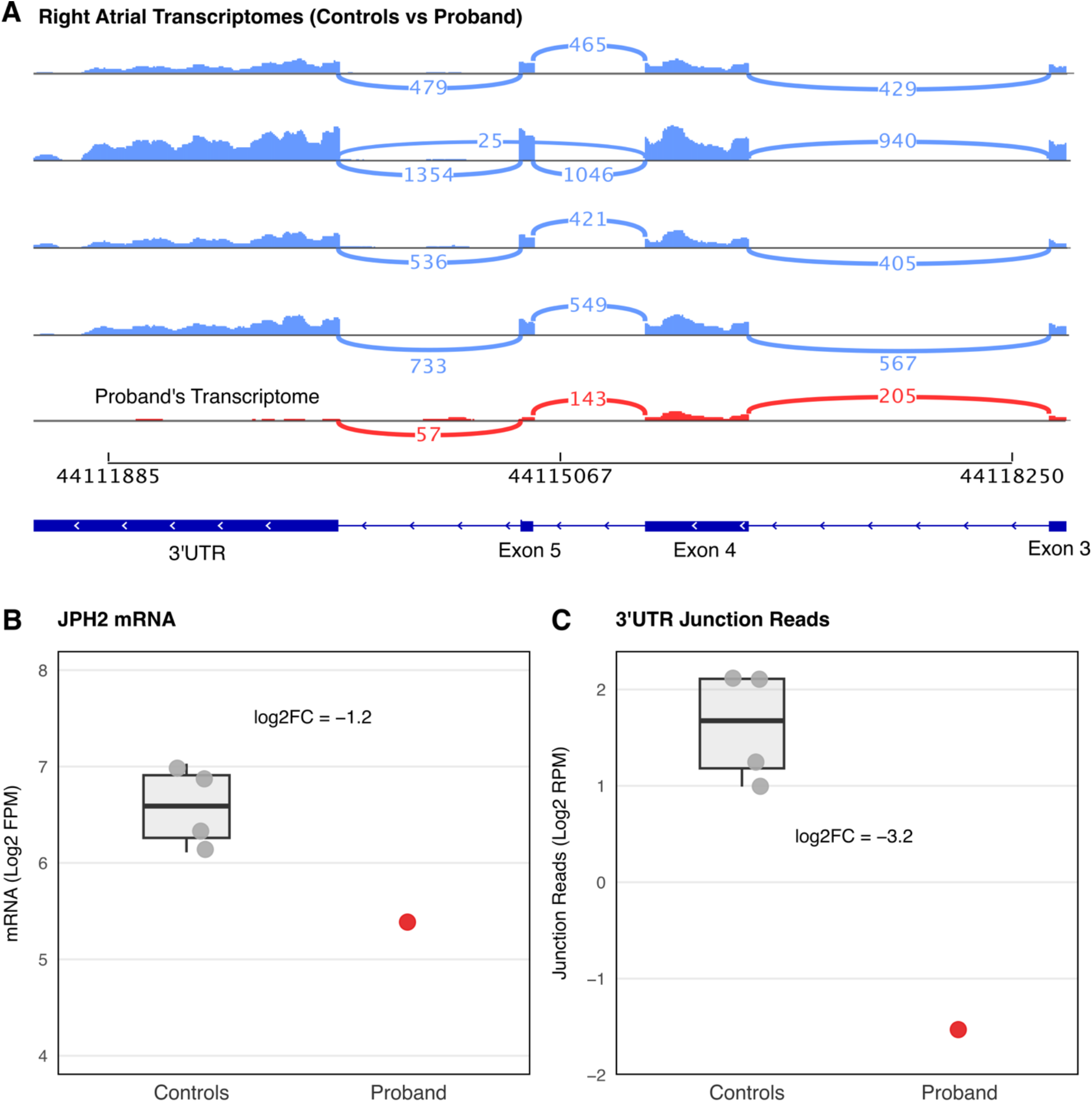
RNAseq analysis using right atrial tissue. RNA sequencing was performed using right atrial tissue from the proband’s explanted heart and four control hearts. **A:** Sashimi plots showing read density and junction reads for *JPH2* (3′UTR to Exon 3, chr20:44,118,250 - 44,111,885). The junction reads were filtered to only show junctions with a minimum of 25 reads. **B:** Overall *JPH2* mRNA expression (log2 FPM - fragments per million) in the five transcriptomes, with the proband’s transcriptome highlighted in red. **C:** Reads spanning to the 3′UTR-Exon 5 junction in the five transcriptomes, with the proband’s transcriptome highlighted in red. Junction reads are normalised to Log2 RPM, with a log2 fold change of -3.2 in the proband’s transcriptome.

## DISCUSSION

Following cardiomyocyte depolarisation, the L-type Ca2+ channels (LTCC) open, permitting Ca2+ influx into the cell. The first wave of Ca2+ influx stimulates the opening of the ryanodine receptor (RyR2) channel, triggering additional Ca2+ release into the cell from the sarcoplasmic reticulum. This process, known as Ca2+-induced Ca2+ release (CICR), is critical for cardiomyocyte contraction. JPH2 is crucial in this context, by forming the junctional membrane complex that directly tethers the transverse tubules of the plasma membrane to the sarcoplasmic reticulum^2,26^. This ensures that the LTCCs on the plasma membrane are maintained in close spatial proximity to the RyR2 channels on the sarcoplasmic reticulum^26^. Due to this structural role, *JPH2* loss-of-function is theorised to compromise Ca2+ handling within cardiomyocytes, precipitating cardiac dysfunction. Alternative non-structural roles for *JPH2* in cardiac physiology are also under investigation, including separate functional roles for the C-terminal and N-terminal JPH2 protein fragments following calpain cleavage^2,27^.

*JPH2* has been recently classified by ClinGen as having a strong association with autosomal recessive DCM^7^. Reported cases of biallelic *JPH2* loss-of-function exhibit a severe paediatric-onset DCM phenotype. Cases include a paediatric patient who died while awaiting a transplant, with a homozygous c.1920dup, p.(Glu641Ter) variant^5^, a paediatric patient who required a heart transplant, with a homozygous c.1282C>T, p.(Gln428Ter) variant^4^, a patient diagnosed with DCM at <10 years of age, with a homozygous c.1426G>T, p.(Glu476Ter) variant^3^, and another paediatric patient diagnosed with DCM, with a homozygous c.575C>A, p.(Ser192Ter) variant^28^. This association between the “genetic knockout” of *JPH2* and DCM is also supported by experimental evidence. For example, cardiac specific *JPH2*-knockdown using RNAi results in acute heart failure with the loss of junctional membrane complexes and reduced excitation-contraction coupling and impaired CICR^29^. Additionally, complete *JPH2* knockout causes embryonic lethality in mice by E10.5-11.5, with aberrant Ca2+ transients in cardiomyocytes from E9.5 embryos^26^.

Although our proband presented at a slightly older age than previously published cases of patients with biallelic *JPH2* loss-of-function, her presentation with paediatric end-stage heart failure is otherwise consistent. Our case therefore adds to the growing literature supporting a role for biallelic *JPH2* loss in the pathogenesis of severe paediatric-onset DCM^7^. Further, both the proband’s parents had normal echocardiograms, consistent with the theory that *JPH2* truncating variants do not cause autosomal dominant DCM.

Crucially, our study illustrates the power of multiomics for elucidating the effects of a structural variant and providing a genetic diagnosis to a family with inconclusive results from clinical exome sequencing. By analysing patient proteomic data we were able to directly pinpoint a defect in JPH2 synthesis, and use this knowledge to identify and resolve a structural variant in the participant’s genome, ultimately providing a genetic diagnosis. This structural variant would not have been detected by exome sequencing due to the inversion breakpoint falling within an intronic region of *JPH2*. Additionally, while RNA sequencing was critical in showing the loss of the 3′UTR from most transcripts, simply performing gene-level differential expression analysis would have only revealed a log2 fold change consistent with monoallelic loss. This highlights the utility of proteomics as a diagnostic adjunct in selected genetically unsolved cases, particularly when disease-relevant tissue is available.

Other proteomics-guided diagnostic approaches have been applied with success in the diagnosis of rare mitochondrial disorders using peripheral blood mononuclear cells (PBMC), fibroblasts, and skeletal muscle biopsy tissue^30^. In our case, an analysis of PBMC-derived proteomic data would have been uninformative due to the low expression of *JPH2* within blood, reflecting the power of having access to patient cardiac tissue.

While obtaining cardiac tissue is not feasible for all patients with DCM, heart transplantation remains the last line of treatment for the most severe cases of paediatric DCM, cases with a higher likelihood of being monogenic. For this subset of severe, likely monogenic cases, proteomics analysis of explanted myocardium will be invaluable for facilitating genetic diagnoses. To the best of our knowledge, we present the first reported use of proteomics from explanted cardiac tissue to provide a genetic diagnosis. Our methodology has broad relevance to patients with other genetically unsolved Mendelian diseases, who might undergo organ transplantation as part of routine clinical management. The ability to use clinically inaccessible tissue for diagnostic purposes should therefore be a relevant consideration for biobanking projects at transplantation centres.

## Data Availability

All data produced in the present study are available upon reasonable request to the authors and following appropriate ethics and legal approval.

## ACKNOWLEDGEMENTS

We sincerely thank the families who participated in this research for their significant contribution to advancing the scientific understanding of rare and inherited genetic diseases.

This research was supported by the Medical Research Future Fund (MRFF) Cardiovascular Health Mission (#2024269), the National Health and Medical Research Council (NHMRC) Synergy Grant (#2035975) and was undertaken with the assistance of resources from the National Computational Infrastructure (NCI Australia), an NCRIS enabled capability supported by the Australian Government.

J.I, D.G.M, E.R.P and S.J.H are supported by NHMRC investigator grants (#2034308, #2009982, #2008376, #2026905). J.I is supported by a National Heart Foundation of Australia Future Leader Fellowship. E.R.P. and S.J.H. are members of The Novo Nordisk Foundation Center for Stem Cell Medicine, reNEW, which is supported by Novo Nordisk Foundation grant no. NNF21CC0073729. The biobanking of patient samples by the Melbourne Children’s Heart Tissue Bank was supported by the Loti and Victor Smorgon Family Foundation and Royal Children’s Hospital Foundation.

Genomic analysis was supported by the Centre for Population Genomics (Garvan Institute of Medical Research and Murdoch Children’s Research Institute) and was funded in part by a NHMRC investigator grant (#2009982) and the MRFF Genomics Health Futures Mission (#2032931). This work also forms part of Australian BioCommons’ GUARDIANS program, which is enabled by NCRIS investment via Bioplatforms Australia. The contents of this published material are solely the responsibility of the authors and do not reflect the views of the Commonwealth of Australia or the NHMRC.

## REFERENCES

1. Landstrom, A. P., Beavers, D. L. & Wehrens, X. H. T. The junctophilin family of proteins: from bench to bedside. Trends Mol Med 20, 353–362 (2014).

2. Hutchison, D. F., Lahiri, S. K., Prins, K. W. & Wehrens, X. H. T. Nonstructural Roles of Junctophilin-2 in the Heart. JACC Basic Transl Sci 10, 101272 (2025).

3. Mehaney, D. A. et al. Molecular analysis of dilated and left ventricular noncompaction cardiomyopathies in Egyptian children. Cardiol Young 32, 295–300 (2022).

4. Vasilescu, C. et al. Genetic Basis of Severe Childhood-Onset Cardiomyopathies. J Am Coll Cardiol 72, 2324–2338 (2018).

5. Jones, E. G. et al. Analysis of enriched rare variants in JPH2-encoded junctophilin-2 among Greater Middle Eastern individuals reveals a novel homozygous variant associated with neonatal dilated cardiomyopathy. Sci Rep 9, 9038 (2019).

6. Parker, L. E., Kramer, R. J., Kaplan, S. & Landstrom, A. P. One gene, two modes of inheritance, four diseases: A systematic review of the cardiac manifestation of pathogenic variants in JPH2-encoded junctophilin-2. Trends Cardiovasc Med 33, 1–10 (2023).

7. Jordan, E. et al. An updated evidence assessment of the genetic causes of dilated cardiomyopathy. medRxiv (2026) doi:10.64898/2026.03.09.26347990.

8. Genes Associated With Hypertrophic Cardiomyopathy: A Reappraisal by the ClinGen Hereditary Cardiovascular Disease Gene Curation Expert Panel. Journal of the American College of Cardiology 85, 727–740 (2025).

9. Collins, R. L. et al. A structural variation reference for medical and population genetics. Nature 581, 444–451 (2020).

10. Pais, L. S. et al. seqr: A web-based analysis and collaboration tool for rare disease genomics. Hum Mutat 43, 698–707 (2022).

11. Laricchia, K. M. et al. Mitochondrial DNA variation across 56,434 individuals in gnomAD. Genome Res 32, 569–582 (2022).

12. Pedersen, B. S. et al. Somalier: rapid relatedness estimation for cancer and germline studies using efficient genome sketches. Genome Med 12, 62 (2020).

13. Payne, A. et al. Readfish enables targeted nanopore sequencing of gigabase-sized genomes. Nat Biotechnol 39, 442–450 (2021).

14. Gamaarachchi, H. et al. Fast nanopore sequencing data analysis with SLOW5. Nat Biotechnol 40, 1026–1029 (2022).

15. Samarakoon, H., Ferguson, J. M., Gamaarachchi, H. & Deveson, I. W. Accelerated nanopore basecalling with SLOW5 data format. Bioinformatics 39, (2023).

16. Kalikar, S., Jain, C., Vasimuddin, M. & Misra, S. Accelerating minimap2 for long-read sequencing applications on modern CPUs. Nat Comput Sci 2, 78–83 (2022).

17. Poplin, R. et al. A universal SNP and small-indel variant caller using deep neural networks. Nat Biotechnol 36, 983–987 (2018).

18. Smolka, M. et al. Detection of mosaic and population-level structural variants with Sniffles2. Nat Biotechnol 42, 1571–1580 (2024).

19. Jiang, T. et al. Long-read-based human genomic structural variation detection with cuteSV. Genome Biol 21, 189 (2020).

20. Martin, M. et al. WhatsHap: fast and accurate read-based phasing. bioRxiv (2016) doi:10.1101/085050.

21. Proudfoot, N. J. Ending the message: poly(A) signals then and now. Genes Dev 25, 1770–1782 (2011).

22. Wieder, N. et al. The role of untranslated region variants in Mendelian disease: a review. Eur J Hum Genet 33, 1096–1105 (2025).

23. Passmore, L. A. & Coller, J. Roles of mRNA poly(A) tails in regulation of eukaryotic gene expression. Nat Rev Mol Cell Biol 23, 93–106 (2022).

24. Hu, J. et al. RBFox2-miR-34a-Jph2 axis contributes to cardiac decompensation during heart failure. Proc Natl Acad Sci U S A 116, 6172–6180 (2019).

25. Bresson, S. & Tollervey, D. Surveillance-ready transcription: nuclear RNA decay as a default fate. Open Biol 8, (2018).

26. Takeshima, H., Komazaki, S., Nishi, M., Iino, M. & Kangawa, K. Junctophilins: a novel family of junctional membrane complex proteins. Mol Cell 6, 11–22 (2000).

27. Weninger, G. et al. Calpain cleavage of Junctophilin-2 generates a spectrum of calcium-dependent cleavage products and DNA-rich NT-fragment domains in cardiomyocytes. Sci Rep 12, 10387 (2022).

28. Janin, A. et al. Molecular Diagnosis of Primary Cardiomyopathy in 231 Unrelated Pediatric Cases by Panel-Based Next-Generation Sequencing: A Major Focus on Five Carriers of Biallelic TNNI3 Pathogenic Variants. Mol Diagn Ther 26, 551–560 (2022).

29. van Oort, R. J. et al. Disrupted junctional membrane complexes and hyperactive ryanodine receptors after acute junctophilin knockdown in mice. Circulation 123, 979–988 (2011).

30. Hock, D. H. et al. Untargeted proteomics enables ultra-rapid variant prioritisation in mitochondrial and other rare diseases. Genome Med 17, 58 (2025).

